# A Pair Formation Model with Recovery: Application to Monkeypox

**DOI:** 10.1101/2022.08.17.22278897

**Authors:** Matthew I Betti, Lauren Farrell, Jane Heffernan

**Affiliations:** Mount Allison University, Sackville, Canada; York University, Toronto, Canada; Centre for Disease Modeling, Toronto, Canada

**Author notes:** MB developed and analyzed the model; LF refined the model; JH analyzed the model and contrbuted to the writing of the manuscipt. The authors have no competing interests.

**Keywords:** Pair Formation, Mathematical Model, Monkeypox

## Abstract

The current global outbreaks of Monkeypox is a unique infectious disease in the way it seems to be transmitting: it has been observed to be highly concentrated in communities of men who have sex with men (MSM) through pair formation, and also provides immunity. This framework of mostly close, prolonged contact spreading a disease that admits immunity after infection is unlike similar infections which either offer little to no immunity post-infection or are lifelong infections. This creates the need for a new model framework that incorporates pair formation structure with recovery. While seemingly a straight forward model, we show how new dynamics arise from the combination of pair formation and recovery that are not present in a standard model with recovery and also not present in a pair formation model without recovery. We see that the combination of these two properties allows for waves of infection that are not seen in a standard SIR model. These dynamics suggest that outbreaks of monkeypox around the world may require special attention from public health. We also derive a reproduction number for this model and estimate the reproduction number of human monkeypox to be ≈ 2.3 using global and Canadian data. The expression derived for *R*_0_ can help estimate key parameters for diseases transmission and public health interventions and compare to equivalent models without pair formation.

**Significance Statement:** With outbreaks of Monkeypox being observed around the world, a modeling framework which takes into account the unique properties of this emerging disease is necessary for understanding the disease and public health mitigation. Monkeypox seems to be unique in that it requires close, prolonged contact with an infected individual in order to spread, but also provides immunity after infection. We develop a model for this situation and show how this differs from simpler models which are currently being used for disease dynamics.

## 1. Introduction

Infections that require close, prolonged contact are more realistically modeled by pair formation models (1). Seuxally transmitted diseases fall within this scope. Many sexually transmitted infections are either treatable, with potential for reinfection (e.g. chlamydia (2, 3), gonorrhea(4, 5)) or are chronic as in the case of HIV (6) and HSV (7). As such, pair formation models have been limited to tracking susceptible-infected or susceptible-infected-susceptible structures.

A seminal model on pair formation was developed in (8). In this study, Kretzschmar *et al*. develop a model for the spread of infection through pair formation for a chronic disease. The model is extended to include two infectious classes as well. Deep insights into the behaviour of the model and its epidemiological interpretations are present in the paper; the basic reproduction number is computed, and it is shown that a model without pair formation can underestimate the overall prevalence of disease in a population (8). The Kretzschmar *et al*. model has also been extended to include long-term and casual partnerships (9, 10). In this model, individuals are allowed to become susceptible again after infection. In this study it is shown that the importance of casual partnerships in spreading infection is dependent on the duration of infection; in short-lived infections, casual partnerships are crucial to spreading infection (9).

In recent times, monkeypox has begun to spread in many global regions (11). Monkeypox, a disease caused by the monkeypox virus, is a relative of the smallpox and the cowpox viruses. The endemic region for monkeypox is historically Central and West Africa (12), first being observed in 1970 (13). The vast majority of monkeypox infections will recover and it is theorized that these individuals gain long-term immunity (14–16). The case fatality ratio of monkeypox is strain dependent with case fatality ratios ranging from 1% to 11% (17) and the more fatal strains having been observed to have human-to-human transmission (17).

Monkeypox transmission requires close, prolonged contact with an infected individual (11). While not directly sexually transmitted, this close, prolonged contact is best modeled by pair formation. Moreover, in the recent international outbreak of monkeypox, observed cases seem to be concentrated in the community of men who have sex with men (18). Cases have have also been tied to international travel (18). As case counts rise above stochastic effects and cases are found outside of sexual encounters(19), the need for a mechanistic model that can capture the routes of transmission and analyze scenarios for disease outcomes is requires.

This creates a unique situation where a disease can be modeled by a pair formation model, but individuals can recover with immunity (20). Moreover, vaccination is possible as it has been observed that inoculation with a smallpox vaccine provides sufficient immunity against monkeypox (21).

In the current study, we develop a framework for a model of pair formation with recovery by extending the model developed in (8). We show how the dynamics of this model differ from a standard *SIR* model, and can lead to multiple waves of infection. We formulate the basic reproduction number for this model that can be used as more information becomes available to better estimate the reproduction of monkeypox within a population; as a need for such an expression has been stated in the literature (22). Lastly, we validate this model versus a standard SIR model by parameterizing both the pair-formation model and a standard SIR model and show that model selection metrics favour the pair-formation model.

We discuss extensions to the model that may prove useful for long-term forecasting of outbreaks, the creation of animal reservoirs, and effective vaccination strategies against further monkeypox outbreaks.

## 2. The Model

### A. Infection through Pair Formation

Current evidence points to Monkeypox being transmitted via prolonged, close contact between individuals; particularly those in the men who have sex with men (MSM) community. Thus, the standard SIR model that assumes instantaneous contacts and a well-mixed population will not suffice here.

A pair formation model structure(8) will form the basis for a model of monkeypox. The model in (8) is insufficient as it assumes lifelong infectivity, therefore we need to add a compartment *R*. The standard model of pair formation, with a susceptible class, *S*, and infectious class *I*, and a recovered class *R*, can be written as

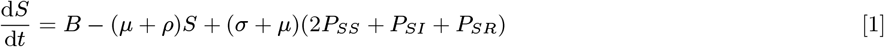

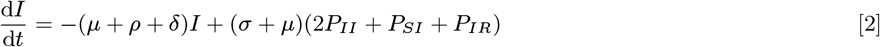

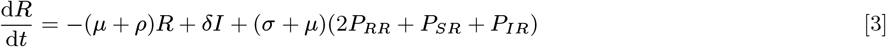

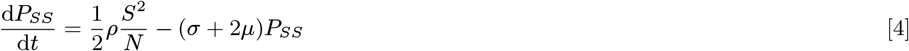

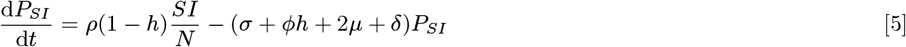

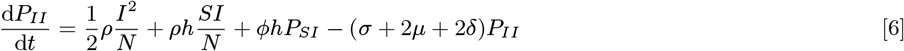

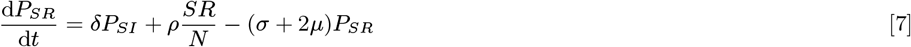

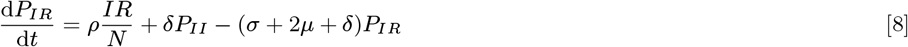

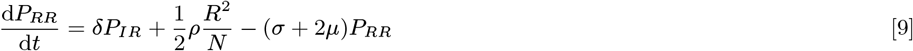

The parameters interpretations are given in Table 1.

**Table 1.** Table of parameters use to explore the model’s qualitative and quantitative features. In Section 5, we estimate model parameters for global and Canadian monkeypox case counts.

| Parameter | Description | Value | Reference |
| --- | --- | --- | --- |
| $B$ | Recruitment into population | $\mu$ | |
| $\mu$ | Removal from population | $(18250 \text{ days})^{-1}$ | (23) |
| $\rho$ | pair formation rate | $(15 \text{ days})^{-1}$ | |
| $\sigma$ | pair dissolution rate | $(1 \text{ day})^{-1}$ | |
| $\delta$ | infection recovery rate | $(30 \text{ days})^{-1}$ | (24) |
| $h$ | probability of transmission | 0.9 | |
| $\phi$ | Contact rate within partnership | 1 | |

The total number of active infections is given by

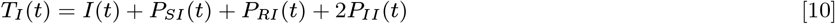

There are some assumptions built-in to this model for the sake of simplicity. The assumptions are

- Pairs are monogamous for the duration of their pairing.
- Groups of three or more cannot be formed.
- If one individual of a pair is removed from the population, the other individual is returned to the single compartment and can form a new pair.
- There is no public health intervention.

### B. SIR with standard incidence

A more common method for modelling close, prolonged contact is through an *SIR* model with standard incidence. The model equations are given by

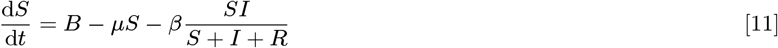

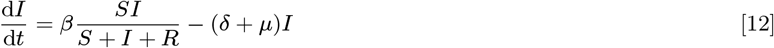

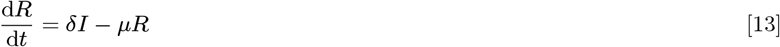

In this model, individuals enter the population through parameter *B*, leave the population through parameter *µ* and recover from infection with immunity at rate *δ*. Infection is passed from an infected individual to a susceptible individual at rate *β*.

The basic reproduction number for this model is simply given by

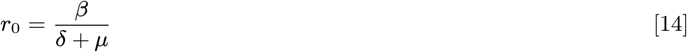

In the following analysis, *β* will be chosen so that *R*_0_ = *r*_0_; while *δ* and *µ* - parameters that are far easier to measure - will be set equal.

## 3. Alternative formulation

Due to the inclusion of recovery, particularly the term *δP*_*II*_ in equation Eq. (8), the model cannot be fully reformulated to remove *P*_*II*_ as in (8). We can however augment the model with the total number of infections

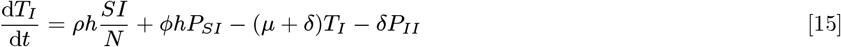

where *T*_*I*_ = *I* + *P*_*SI*_ + 2*P*_*II*_ + *P*_*IR*_.

## 4. Results

### A. The Basic Reproduction Number

The basic SIR pair formation model given by equations Eq. (1) through Eq. (9) is large, but simple in its treatment of disease. Since new infections can only enter the system through the *P*_*II*_ class, the Next Generation Matrix (NGM) approach (25) reduces the system to a matrix system of rank 1, from which we can compute the reproduction number.

We first linearize the system around the disease-free equilibrium given by

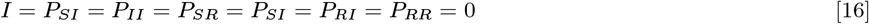

We refactor the linearized system into the standard *F* and *V* matrices, where *F* is the terms related to new infections, and *V* consists of all flux terms between classes and the system has the form

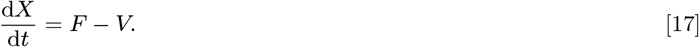

In the case of pair formation, all classes except *P*_*RR*_ participate in the formation of new infections. This leads to an 8 × 8 matrix. The only terms involved in the creation of new infections are

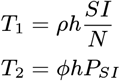

Since both terms appear in class *P*_*II*_, *F* is a sparse matrix of rank 1 and can be written as

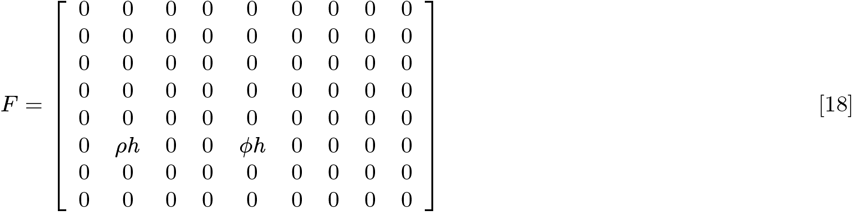

All other terms are relegated to the matrix *V*.

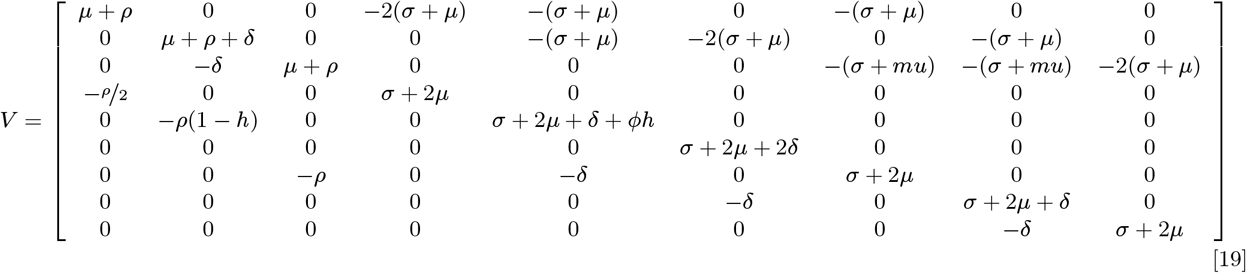

The next generation matrix is then given by

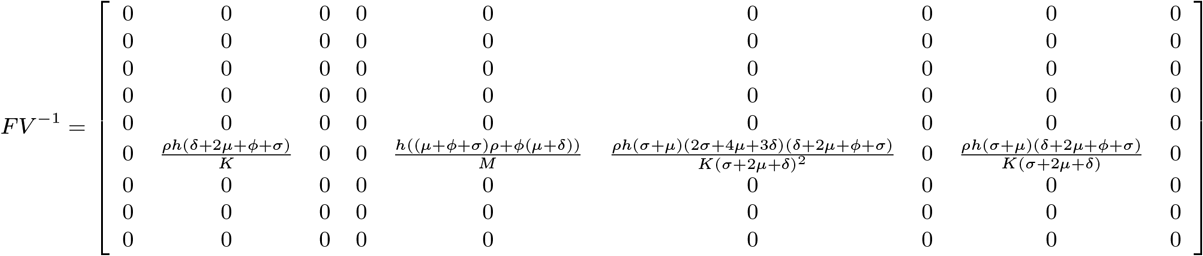

where

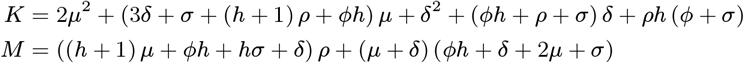

Since *F* is rank 1, the product *FV* ^*−*1^ is also rank 1. This leave a unique, non-zero eigenvalue for the next generation matrix. By definition, this eigenvalue can be interpreted as *R*_0_. It is given by

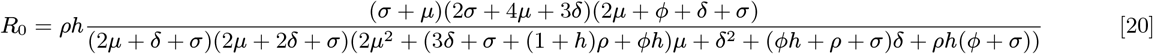

This equation is numerically validated in Figure 1. While the expression is closed and can be used for monitoring, forecasting and policy purposes, the nature of the NGM approach leaves this particular expression difficult to interpret.

**Fig. 1.**
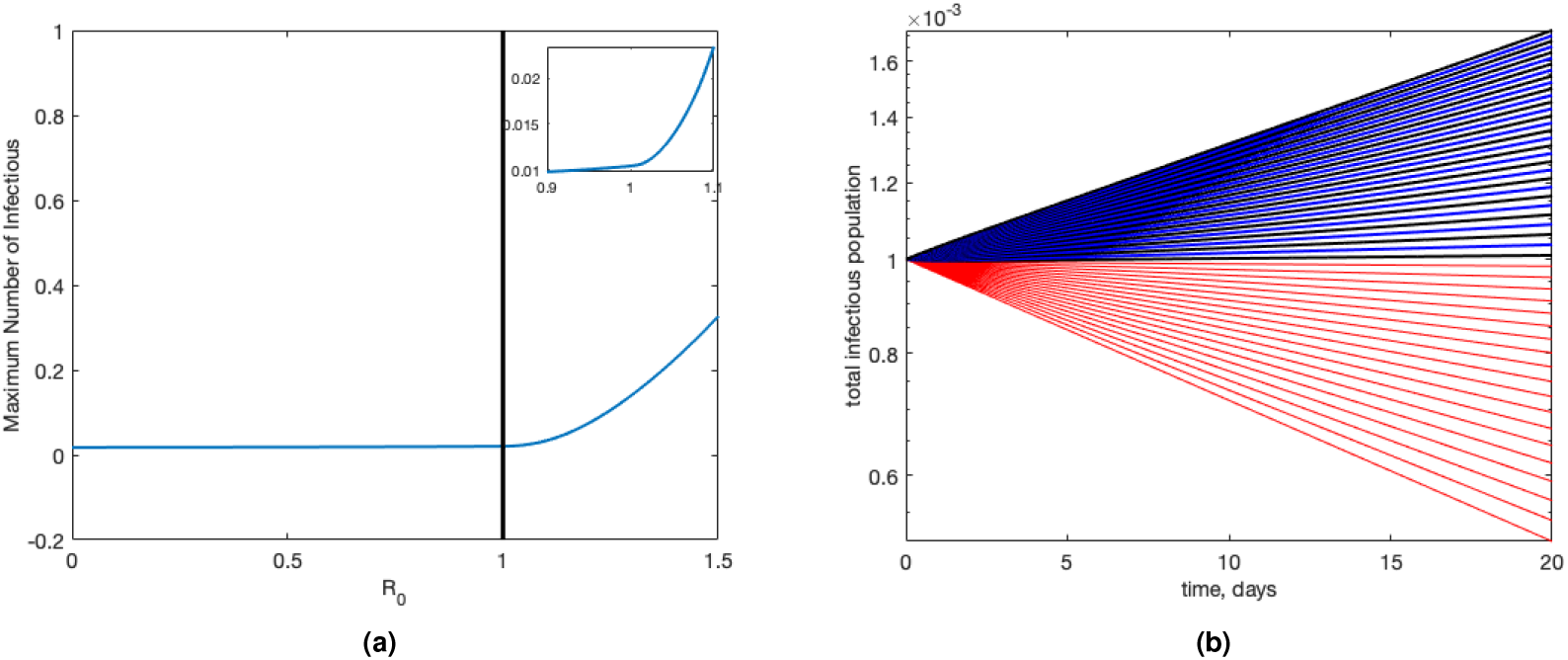
Numerical confirmation of *R*_0_ (equation Eq. (20)), visualized in two ways. We start all simulations with *I*(0) = 1 *×* 10^*−*3^. Panel (a): The maximum number of infections for the model given by equations Eq. (1) through Eq. (9) as a function of *R*_0_. We see that the expression for *R*_0_ provided by equation Eq. (20) is verified numerically. Panel (b): Infection curves for different values of *R*_0_. Red curves represent where *R*_0_ *<* 1 and black/blue curves are scenarios where *R*_0_ *>* 1. Colours alternate between black and blue for clarity.

### B. Alternative Reproduction Number

Recreating the next generation matrix with all new infections entering the system through equation Eq. (15), and infectious individuals transitioning through the classes *I, P*_*SI*_, *P*_*II*_, and *P*_*IR*_ allows an alternative formulation of the basic reproduction number that is more readily comparable to (8). This is given by

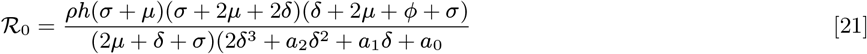

where

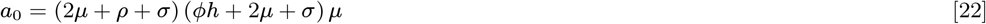

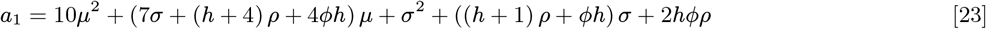

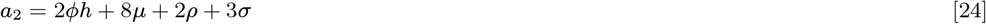

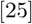

When *δ* = 0, this formulation agrees exactly with the basic reproduction number given in (8). Unlike the reproduction number for the model without recovery(8), the denominator here cannot be nicely factored.

We note here that *ℛ*_0_ and *ℛ*_0_ have the same threshold value, as expected.

### C. Average Number of Partners during Infection

Using the ansatz that

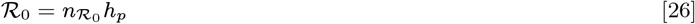

in other words, the basic reproduction number is the product of the number of partnerships formed in one infectious lifetime, *n*, and the probability of infection per partnership, *h*_*p*_, we can use either of our formulations to estimate the average number of partnerships of one infected individual in a completely susceptible population.

At the beginning of an outbreak, we may assume that the entire population is susceptible and divided between classes *S* and *P*_*SS*_. Importantly, *P*_*SI*_ = *P*_*II*_ = *P*_*IR*_ = *R* = 0. This means that the only partnerships that can form at the beginning of an outbreak which involve a susceptible individual are *P*_*SI*_. In this case, setting *h* = 1 would imply *h*_*p*_ = 1. Thus, 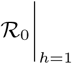, or 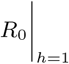, gives the number of partnerships during one infectious lifetime. Mathematically we see this realized as

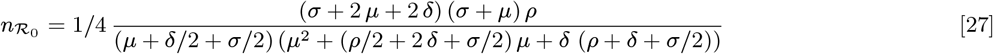

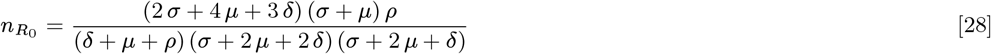

Again, when *δ* = 0, equation Eq. (27) corresponds exactly to *n* as presented in (8).

### D. The limit *σ → ∞*

The limit as *σ* approaches infinity corresponds to the case when pairings become transient contacts. In this case the model, as well as *R*_0_ given in equation Eq. (21), reduce to a simple SIR model with the basic reproduction number being given as

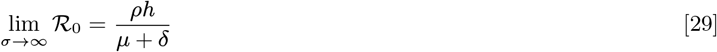

This limit is confirmed numerically in figure 2

**Fig. 2.**
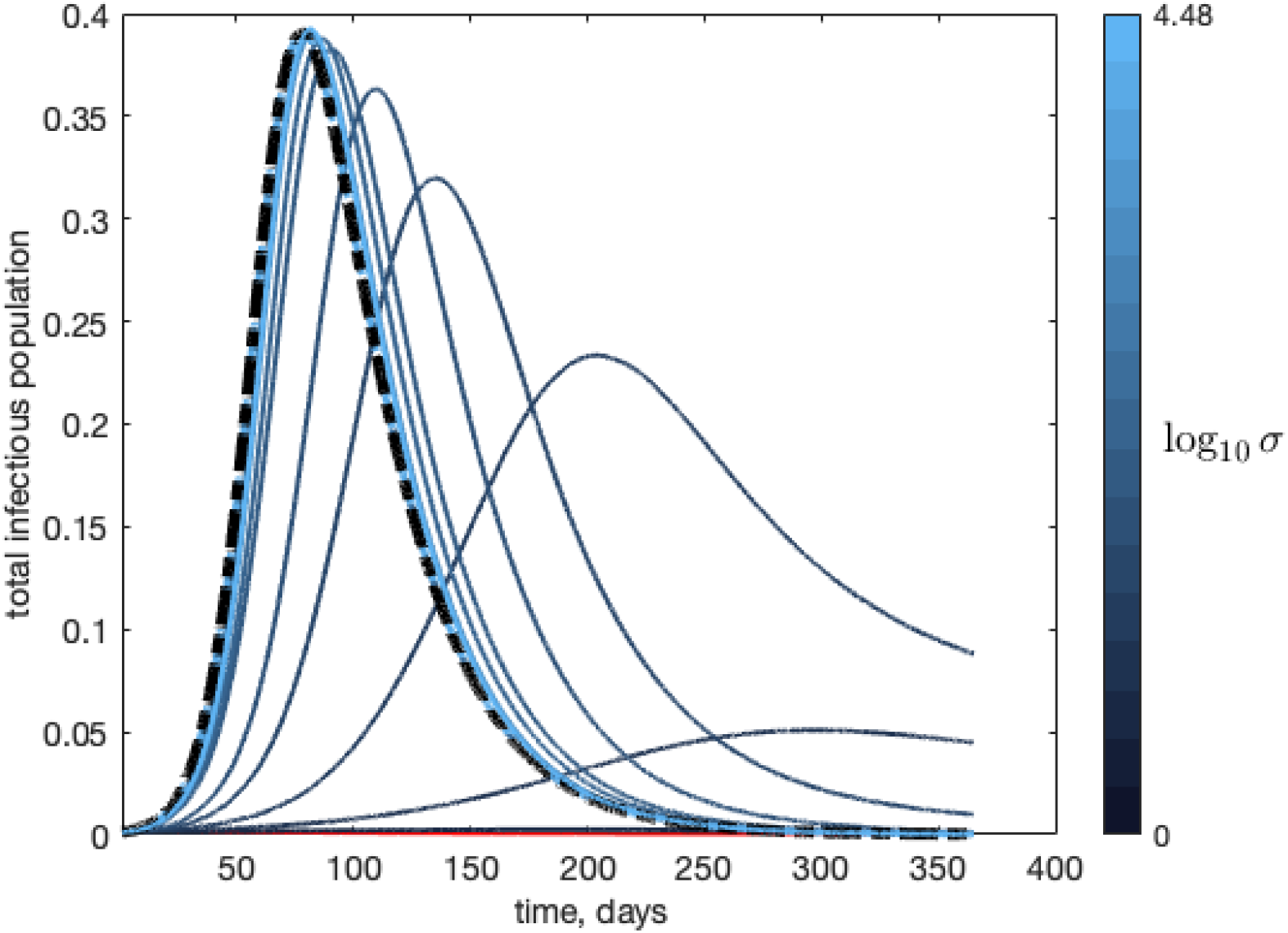
As *σ → ∞*, the pair model approaches the standard SIR model with mass action. The black broken line shows the total number of infections for the SIR model and the solid lines show the total infections over time as *σ* gets large.

### E. Simulations

All simulations are normalized to *R*_0_ in equation Eq. (20) to keep the results as general as possible. For human monkeypox, early estimates of *R*_0_ range between 1.1 and 1.26 (26), and it was hypothesized that human-human transmission before the current outbreaks had a reproduction number less than 1 (27). Therefore, we focus our study on values of *R*_0_ close to 1, and approaching 2 as the upper bound of estimates seems to be around 2(28). There are some parameters that were sourced from the literature on monkeypox. Other parameter values are provided for posterity, although they have no inherent value and are chosen for simplicity and to acquire an *R*_0_ in the correct range. All parameter values are given in Table **??**.

The parameter values provided in Table 1, particularly *ρ* and *σ*, define a scenario where most partnerships are casual and short. When most partnerships are predominantly long, *R*_0_ *<* 1 and the population sees little risk of an epidemic.

With a formulation of *R*_0_, we can look at infection curves for various values of *R*_0_. We normalize our initial susceptible population to *S*_0_ = 1 and look at the infectious proportion of the population, (*I*+*P*_*SI*_ +2*P*_*II*_ +*P*_*IR*_)*/*(*S*+*I*+*R*).

Figure 3 shows that if *R*_0_ is sufficiently large, we can expect a large initial outbreak that will burn itself out relatively quickly, while smaller values of *R*_0_ will lead to a longer but less severe outbreaks, in terms of peak magnitude and width of the infection curve.

**Fig. 3.**
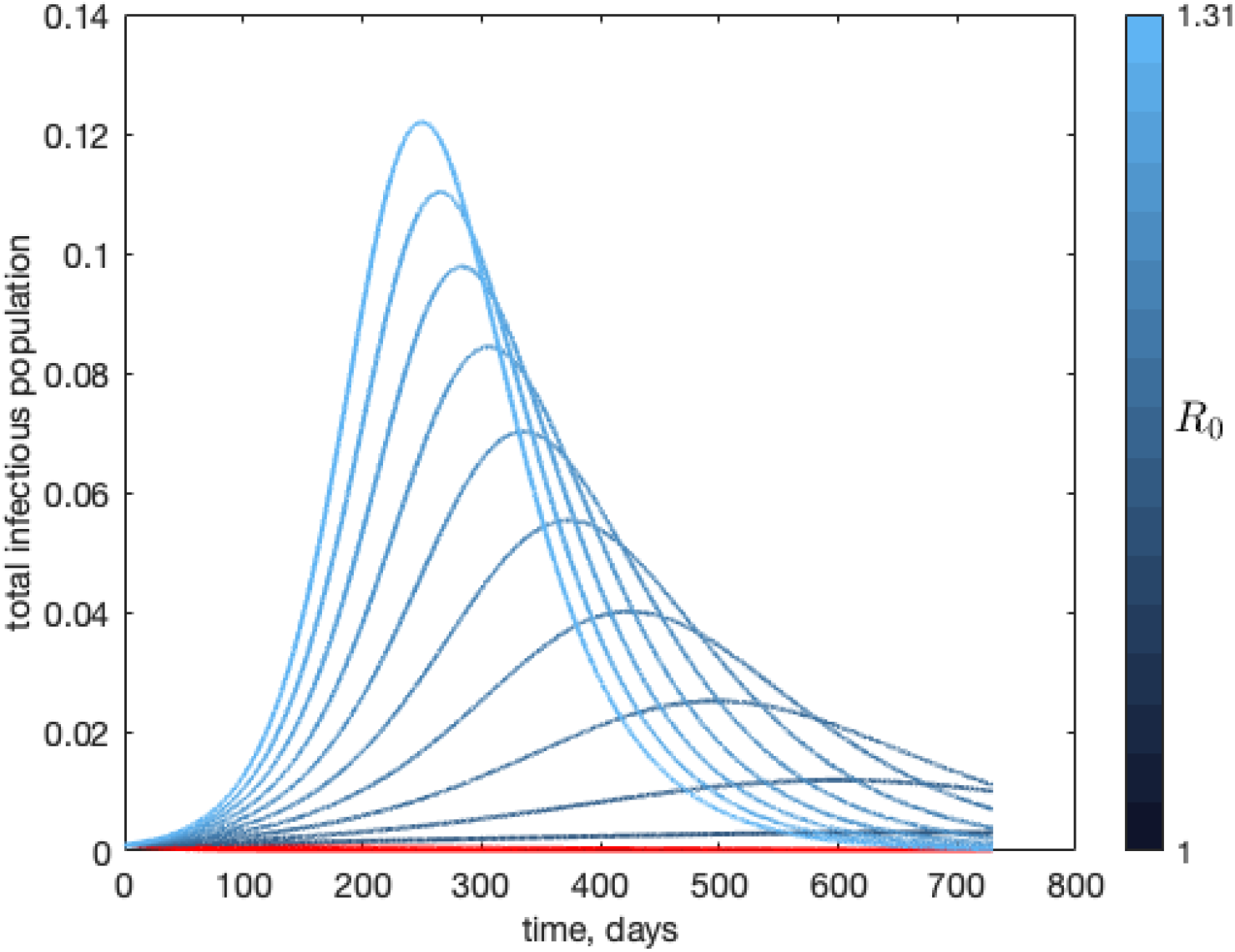
The different outcomes of an outbreak as a function of *R*_0_. The total infectious population, *I* + *P*_*SI*_ + 2*P*_*II*_ + *P*_*IR*_, is shown for a range of *R*_0_, from 1 to 1.31.

Interestingly, as shown in Figure 4, waves of infection can be result from Model given by equations Eq. (1) - Eq. (9). The waves are driven by the introduction of new individuals to the population, through parameter *B*, and through the ability to dissolve old and develop new pairs. Multiple waves of infection that are separated by a period of relative inactivity result. Figure 4 also shows that a less severe first outbreak can lead to a much more severe second outbreak if no public health interventions occur. This can occur because the classes *P*_*II*_ and *P*_*IR*_ can act as a reservoir of infection.

**Fig. 4.**
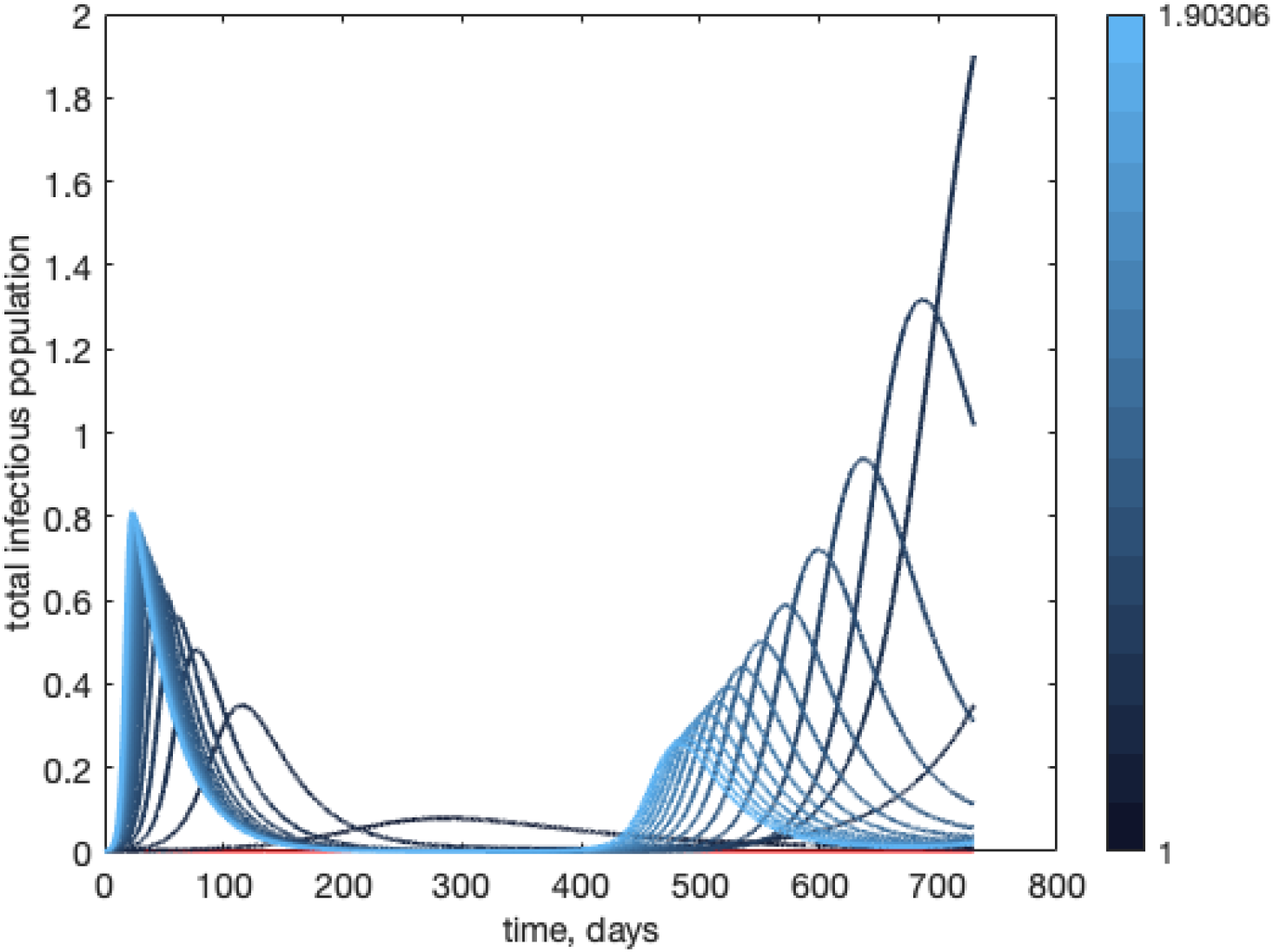
Multiple waves of infection. The total infectious population, *I* + *P*_*SI*_ + 2*P*_*II*_ + *P*_*IR*_, is shown for a range of *R*_0_. Pair formation coupled with the ability to recover from infection leads to multi-wave dynamics.

Figure 5 compares the pair formation model to the standard *SIR* model with normalized incidence (equations Eq. (11) through Eq. (13)) for *R*_0_ = 1.9. Here, we see that the multiple waves of infection are only possible due to the pair formation dynamics, and that the standard *SIR* model significantly underestimates the prevalence in the population while overestimating the amount of time an outbreak lasts.

**Fig. 5.**
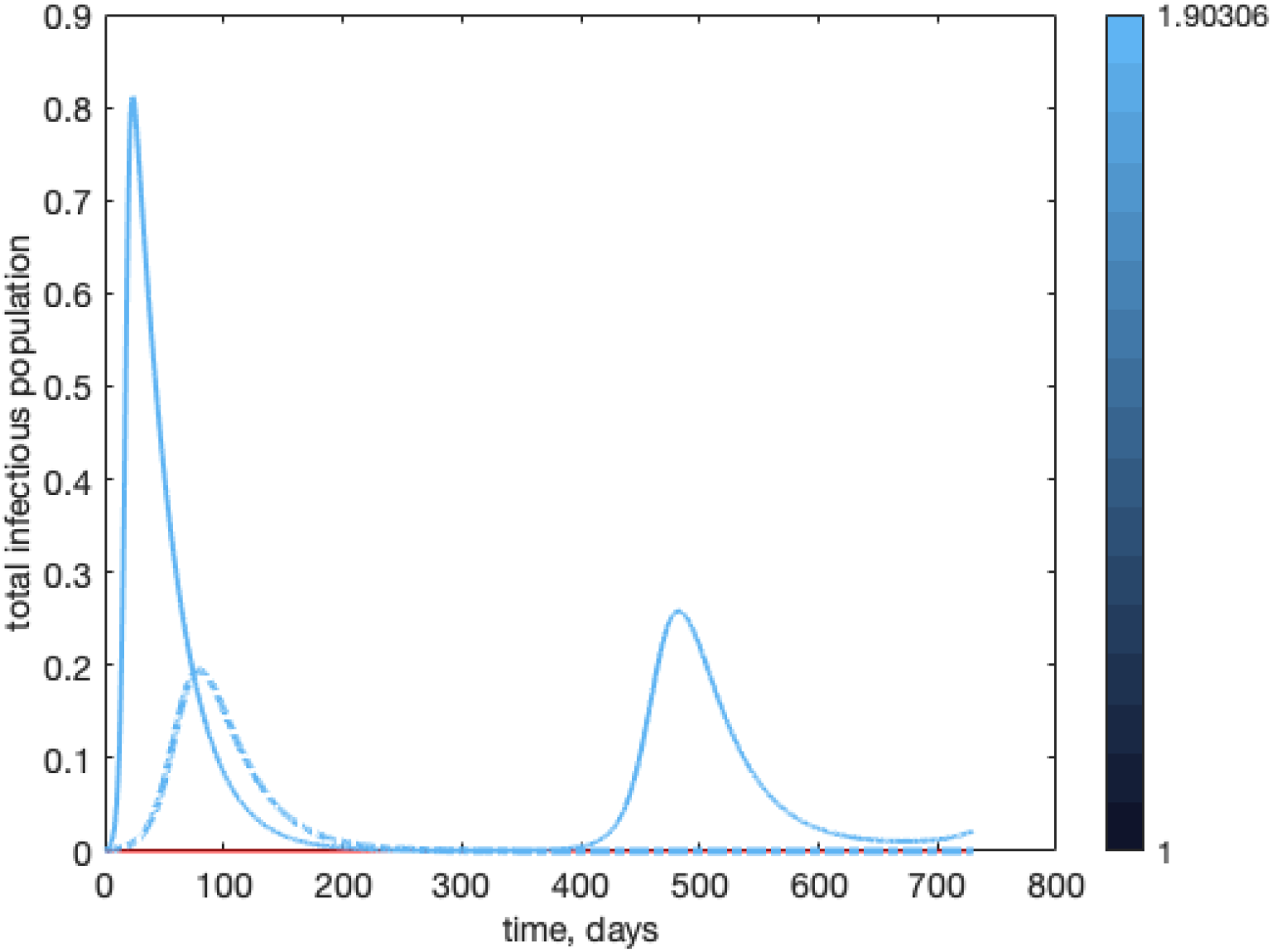
A comparison of the pair formation model (equations Eq. (1) to Eq. (9)) with an *SIR* model with standard incidence (equations Eq. (11) to Eq. (13)). Parameters for both models are chosen so that *R*_0_ = 1.9 for both models.

## 5. Model Validation with Canadian and Global Monkeypox Data

So far, we have shown that a disease that requires finitely long close contact - as opposed to the instantaneous contact assumed by a standard SIR model - can create qualitatively different results with multiple waves of infection. We now turn to using the model to estimate parameters for a population dealing with human Monkeypox.

We fit equations Eq. (1) to Eq. (9) to human Monkeypox data for Canada as reported by the government of Canada (29); the same source provides us with global numbers. We use a standard least squares non-linear regression on the cumulative case counts and the new cases per day. We also fit the data to a standard SIR model.

For our fitting we fix *µ* = *B* = 0 as our time period is much shorter than the demographic time scales, we also fix *δ* = ^1^*/*_30_ as this parameter is fairly well-established in the literature. In order to account for the fluctuations in reporting rate and the true start time of the epidemic, we allow the initial conditions *S*(0), *I*(0), and -where applicable - *P*_*SS*_ (0) to be fit as well.

The parameter estimates along with the Akaike and Bayesian Information Criteria are given in table 2 and the pair formation model fit shown in Figures 6 and 7.

**Table 2.** **Table of fitted parameters for Canadian and global data, model selection parameters and basic reproduction number. *Basic reproduction numbers are computed using equations** Eq. (29), Eq. (20), **and** Eq. (21). **Data is from the Canadian government (29)**.

| Parameter | Canada |  | World |  | Fixed |
| --- | --- | --- | --- | --- | --- |
|  | SIR model | Pair Formation Model | SIR model | Pair Formation Model |  |
| $\delta$ | $1/30$ | $1/30$ | $1/30$ | $1/30$ | × |
| $\mu$ | 0 | 0 | 0 | 0 | × |
| $S(0)$ | 853.83 | 902.56 | 90681 | 29709 | |
| $I(0)$ | 1 | 16.97 | 1 | 0.284 | |
| $P_{SS}(0)$ | N/A | 145.96 | N/A | 14350 | |
| $\rho$ | N/A | 0.30 | N/A | 0.58 | |
| $\phi$ | N/A | 0.91 | N/A | $7.2 \times 10^{-5}$ | |
| $h$ | N/A | 0.395 | N/A | 0.89 | |
| $\sigma$ | N/A | 11.17 | N/A | 0.1722 | |
| $\beta$ | $1.68 \times 10^{-4}$ | N/A | $9.78 \times 10^{-7}$ | N/A | |
| $R_0$ | 4.3* | $\mathcal{R}_0 = 2.7^*, R_0 = 1.45^*$ | 2.66* | $\mathcal{R}_0 = 2.31^*, R_0 = 1.45^*$ | |
| AIC | 1226 | 991 | 2354 | 2324 |  |
| BIC | 1235 | 1013 | 2364 | 2347 |  |

**Fig. 6.**
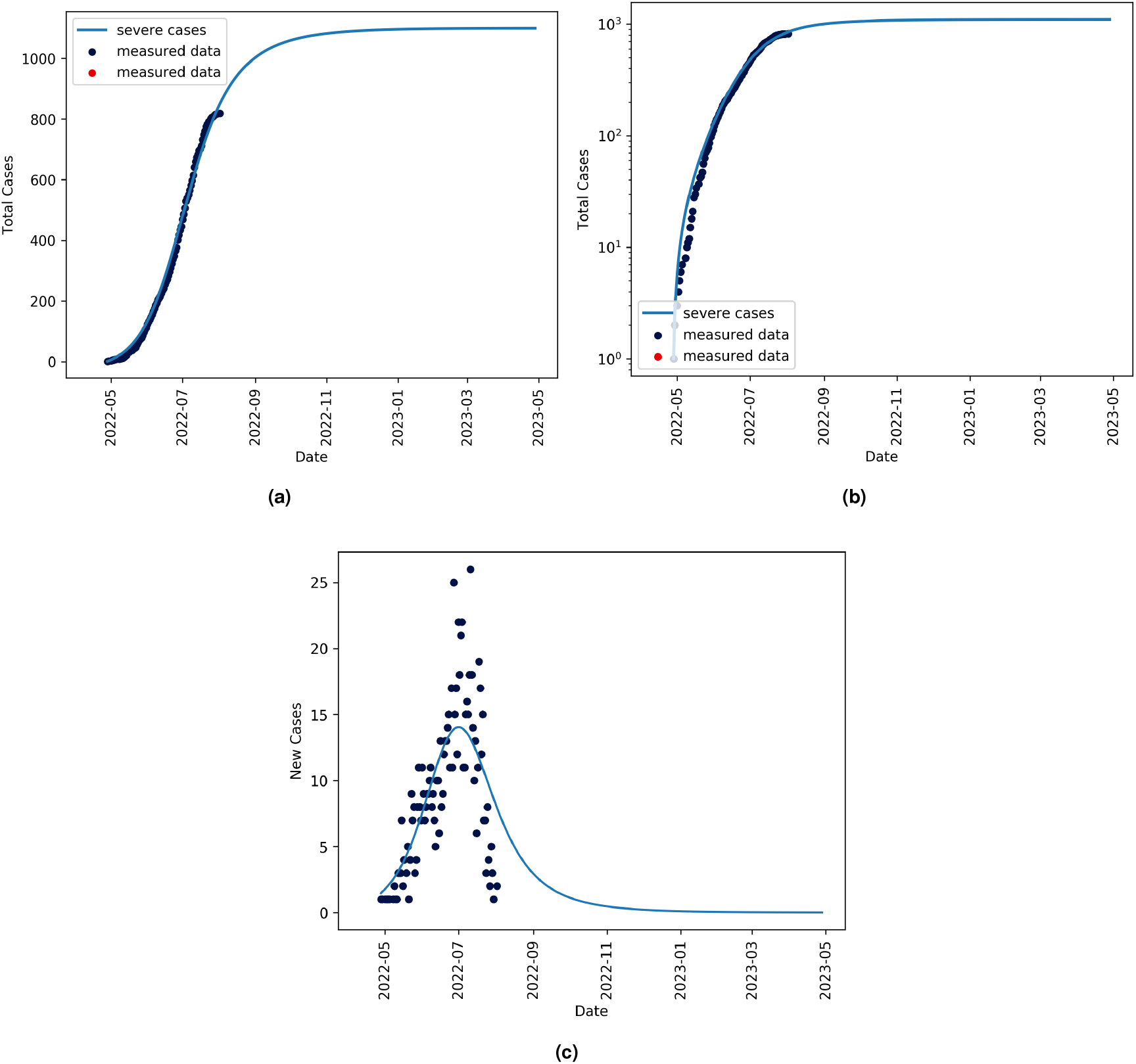
Curves for cumulative infections in Canada on a linear scale (a) and log scale (b) for clarity. Panel (c) shows the new cases per day. Canadian data is taken from (29) and fits are generated using equations Eq. (1) to Eq. (9).

**Fig. 7.**
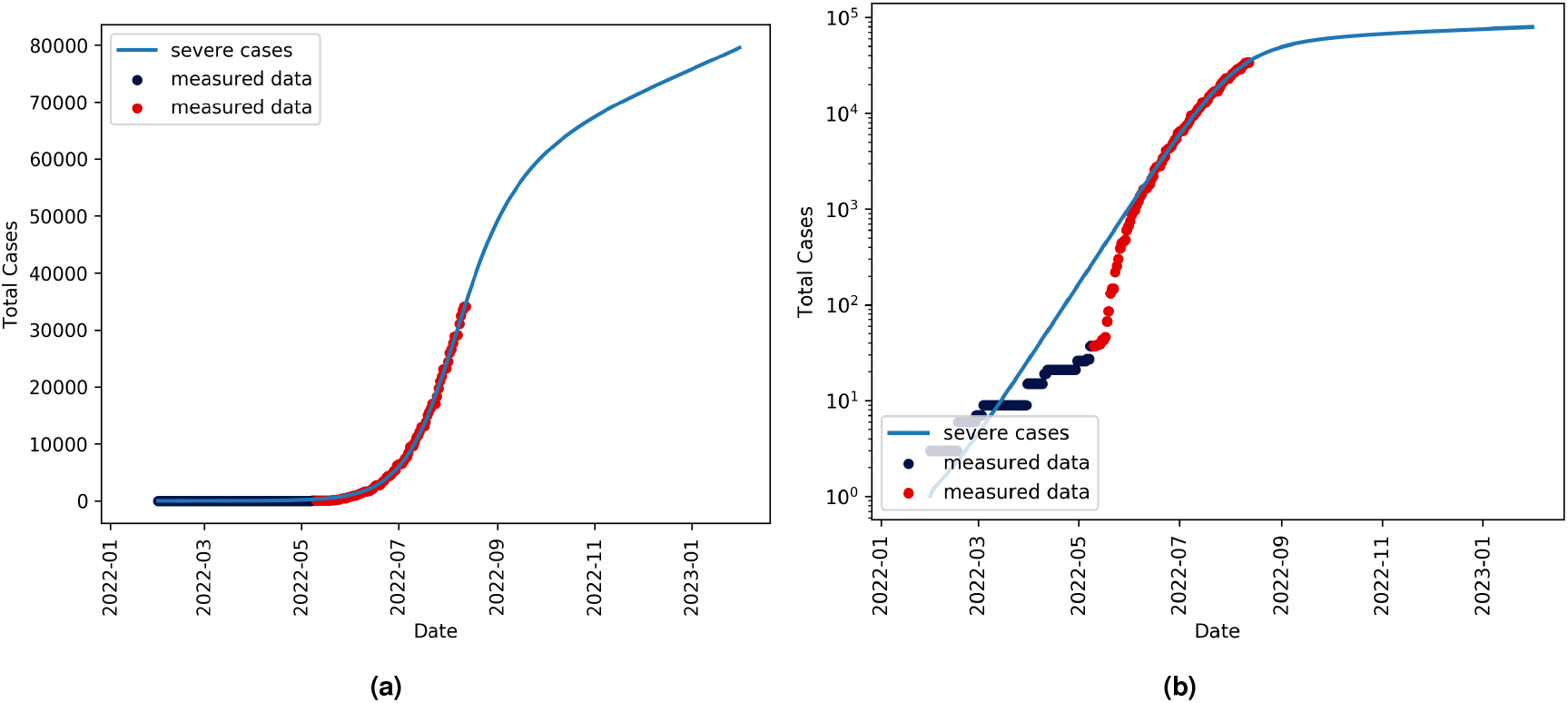
Curves for cumulative infections globally on a linear scale (a) and log scale (b) for clarity. World data is taken from (29) and fits are generated using equations Eq. (1) to Eq. (9).

## 6. Discussion

The model describes the novel scenario under which an infection is predominantly spread through close, prolonged contact but allows for recovery with immunity. This scenario has been overlooked in the literature as most STIs do not bestow immunity to the infected. Evidence to date shows that monkeypox is novel in that it fits this scenario.

Our formulation of *R*_0_ in equation Eq. (20) can be used as more data becomes available to develop estimates of either *R*_0_ or possible contact rates, *ρ* and *σ*, between individuals in any population.

It is interesting to note that our expression for *R*_0_ and the alternative expression for *R*_0_ cannot be readily factored. This is due to the complexity of relationships between classes. As there are certain relationships that can be formed in this model where transmission is impossible, namely *P*_*IR*_, the probability of transmission per partnership is intimately tied to pair formation. These dynamics prevent the two processes from being decoupled except for in limiting cases, like *h* = 1 or *δ* = 0.

Having two expressions for the basic reproduction number leads to questions of which has more real-world applicability. We present both formulations for posterity. Questions of which to use and when are difficult to answer without more data on monkeypox, or experimental estimates of the parameters that make up the basic reproduction number. It is likely that each finds validity in a different parameter regime, and this will be the subject of further study. For instance, the expression for *R*_0_ has the benefit of reducing exactly to the basic reproduction number for the pair formation model without recovery. On the other hand, *R*_0_ leads to a factored and more readily interpretable expression for 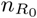.

Our results show that the pair formation *SIR* model differs in dynamics from a typical *SIR* model in that individuals in partnerships with immune individuals can create a reservoir which can lead to a future secondary outbreak. The severity of the secondary outbreak is inversely proportional to the severity of the initial outbreak. Of course, if pair formations are increasingly frequent, *ρ→ ∞* and *σ→ ∞* the dynamics of this model approach those of a standard SIR model.

In terms of public health measures, this means that resources should be put toward monitoring and suppressing spread even after the initial wave of cases seems to have subsided as a lack of public health measures during this time can lead to a secondary epidemic.

More generally, the limit results show that this model is essentially a refinement on the standard SIR model. This model may be more useful for any infectious disease that requires prolonged exposure, or where forecasting accurate quantitative infection curves is of greater importance. The values of *σ* and *ρ* are also more readily interpretable and measureable than the often vague and nebulous contact rate given in a standard mass action model. This can lead to more accurate estimates of *R*_0_.

While currently data on this new monkeypox epidemic is relatively sparse, the model can nonetheless be developed and extended to allow for a wide range of scenarios. While the number of compartments grows quite quickly with each new complexity, the number of parameters does not grow as fast allowing for robust usage even with only moderate quality data.

We use available data for Canada to fit the model. Interestingly, despite having more than twice as many parameters, the model selection criteria used - the AIC and BIC - both confirm that the increase in likelihood outweighs the additional complexity when compared to a standard SIR model.

The parameters estimated can help inform target criteria for vaccination of a high-risk group. For instance, our model predicts *ρ* to have a value of 0.30 which translates to roughly as two close contacts per week and *σ* suggests that close contact be defined as approximately 2 hours of contact. Of course, with better data that is more targeted, the estimates would be far better. Also of note is that the reproduction number between the SIR and pair-formation model is in closer agreement when fitting global data. This allows us to state that the estimated basic reproduction number of human monkeypox to be ≈ 2.31 − 2.66.

One of the hardest parameters to estimate - and why confidence intervals for validation cannot be easily stated - is the effective susceptible population, *S*(0). Since this model is assuming a homogeneous, pair-forming population (i.e. men who have sex with men) who are particularly high-risk it is unclear what the total susceptible population. By allowing this value to be fit, we get an idea of the possible final size of an outbreak at the expense of ill-defined confidence intervals. Extending this model to include other key demographic populations would allow this initial condition to be set and allow for a far more robust fitting of data. Of course, this comes at the expense of a more complex model.

Figure 7 highlights the qualitative dynamics of the pair formation model that are impossible in the standard SIR model. We see that after a period of sharp growth, we see sustained slow growth of cases over time. This is in contrast to a standard SIR model which will plateau.

The model can be extended to incorporate a high-risk and low-risk population to determine potential for spillover from a high-risk population into a larger low-risk population.

In the case of monkeypox, it is known that the virus can and does survive in a variety of animal hosts (30). While these animal reservoirs are currently localized to West and Central Africa (12), the global spread of monkeypox creates the risk that animal reservoirs, particularly in rodents and pets (31), can be created in other parts of the world. The model presented here can be augmented to include an animal reservoir to assess risk of such a scenario occurring and explore preventative measures.

## Data Availability

All data produced are available online at https://health-infobase.canada.ca/monkeypox/

## ACKNOWLEDGMENTS

Please include your acknowledgments here, set in a single paragraph. Please do not include any acknowledgments in the Supporting Information, or anywhere else in the manuscript.

